# Risk factors for severe COVID-19 differ by age: a retrospective study of hospitalized adults

**DOI:** 10.1101/2022.02.02.22270287

**Authors:** Sevda Molani, Patricia V. Hernandez, Ryan T. Roper, Venkata R. Duvvuri, Andrew M. Baumgartner, Jason D. Goldman, Nilüfer Ertekin-Taner, Cory C. Funk, Nathan D. Price, Noa Rappaport, Jennifer J. Hadlock

**Affiliations:** Institute for Systems Biology, 401 Terry Ave N, Seattle, WA, 98109, USA; Swedish Center for Research and Innovation, Seattle, WA 98109, USA; Providence St. Joseph Health, Renton, WA 98057, USA; Division of Allergy & Infectious Diseases, University of Washington, Seattle, WA 98109, USA; Department of Neuroscience, Department of Neurology, Mayo Clinic Jacksonville, Jacksonville, FL 32224, USA; Longevity, a division of Thorne HealthTech, New York, USA; Institute for Systems Biology, 401 Terry Ave N, Seattle, WA, 98109, USA

**Keywords:** COVID-19, Risk, Age Groups, Decision Support Systems, Clinical, Electronic Health Records, Machine Learning

## Abstract

**Background:** Risk stratification for hospitalized adults with COVID-19 is essential to inform decisions for individual patients and allocation of potentially scarce resources. So far, risk models for severe COVID outcomes have included age but have not been optimized to best serve the needs of either older or younger adults. Additionally, existing risk models have been limited to either small sample sizes, or modeling mortality over an entire hospital admission. Further, previous models were developed on data from early in the pandemic, before improvements in COVID-19 treatment, the SARS-CoV-2 delta variant, and vaccination. There remains a need for early, accurate identification of patients who may need invasive mechanical ventilation (IMV) or die, considering multiple time horizons.

**Methods:** This retrospective study analyzed data from 6,906 hospitalized adults with COVID-19 from a community health system with 51 hospitals and 1085 clinics across five states in the western United States. Risk models were developed to predict mechanical ventilation illness or death across one to 56 days of hospitalization, using clinical data collected available within the first hour after either admission with COVID-19 or a first positive SARS-CoV-2 test. The relative importance of predictive risk factors features for all models was determined using Shapley additive explanations.

**Findings:** The percentage of patients who required mechanical ventilation or died within seven days of admission to the hospital due to COVID-19 was 10.82%. For the seven-day interval, models for age ≥ 18 and < 50 years reached AUROC 0.80 (95% CI: 0.70-0.89) and models for age ≥ 50 years reached AUROC 0.83 (95% CI: 0.79-0.88). Models revealed differences in the statistical significance and relative predictive value of risk factors between older and younger patients, including age, BMI, vital signs, and laboratory results. In addition, sex and chronic comorbidities had lower predictive value than vital signs and laboratory results.

**Interpretation:** For hospitalized adults, baseline data that is readily available within one hour after hospital admission or a first positive inpatient SARS-CoV-2 test can predict critical illness within one day, and up to 56 days later. Further, the relative importance of risk factors differs between older and younger patients.

## Introduction

The number of global confirmed cases with severe acute respiratory syndrome Coronavirus 2 (SARS-CoV-2) infection has surpassed 257 million as of December 10, 2021, with over 5 million reported deaths.^1^ Although the majority of patients infected by SARS-CoV-2 present with mild symptoms, studies reported that 20% get hospitalized and 5% of patients with Coronavirus disease 2019 (COVID-19) become critically ill.^2,3^ From early on of the pandemic, both age and chronic comorbidities have been reported as a significant risk factor for poor outcomes^4^, and evidence supports increased risk with hypertension, diabetes, chronic obstructive pulmonary disease, chronic renal disease, and cardiovascular conditions.^4–6^ Although young patients have a lower prevalence of comorbidities than aging patients, the relative risk of fatal outcome in young patients with hypertension, diabetes and cardiovascular diseases has been shown to be higher than in elderly patients.^7^ In addition, some studies show patient population tends to be younger with the emergence of delta as the variant of concern in U.S. with regional proportions being greater than 99% as of November 2021.^8^ Assessing risk for severe COVID-19 in specific age groups is complicated by both the heterogeneity of clinical presentation and age-related differences in the prevalence of chronic multimorbidities. A deeper understanding of risk factors for COVID-19 severity among different age subpopulations is needed, as well as practical, explainable risk stratification for bedside clinical decision support, research stewardship, and advancing our biomedical understanding of SARS-CoV-2.

Several studies have described successful development of machine learning models to predict COVID-19 outcomes in hospitalized patients.^9–17^ Further, explainable models can also inform care decisions by showing which factors lead a specific individual patient to be at risk for severe outcomes, and can also help show which variables are most important at the population level, suggesting areas for further research investigation.^18^ However, existing studies have several limitations; 1) most are based on small sample sizes from academic centers, 2) higher incidence of severe outcomes in hospitalized cohorts than are typically observed with current treatments, 3) reliance on laboratory tests that are not routinely administered to all patients, 4) lack of investigation of differences in risk factors between younger and older hospitalized patients, and 5) marginal model performance for either of age groups.^11^ In this study, we report age-stratified machine-learning models to predict the severity of COVID-19 progression within one, seven, 14, 28, 56 days of the time of hospital admission. The models are developed from data for 6,906 patients from community hospitals across a large geographic area in the western United States over five months after the delta variant became predominant.

## Research in Context

### Evidence before this study

We searched google scholar, PubMed and its associated LitCovid repository for publications in English from database inception until November 15, 2021, using the terms “COVID-19”, “critical illness”, “mortality”, “hospitalized”, “risk score” and “prediction”. The studies we identified generally focused on small cohorts or limited geographic regions and time periods with higher incidence of severe outcomes in hospitalized patients. Earlier papers with more inclusive cohorts were limited to predicting mortality, and we were unable to find papers which developed risk models for current COVID-19 standard of care and Delta variant predominance. None of the studies were optimized to model critical illness or mortality in different age groups or investigate differences between risk factors in each age-group. Lack of age-stratified models has also caused lower performance of general models for younger and older patients.

### Added value of the study

In this study, we analyzed the data from a large cohort across five states in western United States from June 31, 2021 to November 15, 2021, looking at hospitalized patients who were not already on invasive mechanical ventilation. We investigated risk factors for the need for invasive mechanical ventilation or death by developing linear and non-linear complex age-stratified models to accurately predict for time horizons of one to 56 days. Models were based on readily available data within one hour after either hospital admission with COVID-19 or a first positive inpatient SARS-CoV-2 test. In addition, we analyzed how risk factors differ by age.

### Implications of all the available evidence

We present a set of age-stratified COVID-19 critical illness and mortality prediction analyses, derived from large population of patients over a wide geographic region, in the context of current standard of care for COVID-19 and delta-variant predominance. The cohort and analytical methods that we used resulted in prediction models with strong performance, considering there is now lower severe COVID-19 incidence than early in the pandemic. Based on previous studies and linear statistical analyses on the population in this study, comorbidities such as diabetes, hypertension, cardiac disease, chronic lung and kidney disease, and immunosuppression, along with factors including higher age, being male or African American and Hispanic have been associated with higher mortality in patients with COVID-19. However, our study suggests those chronic historical factors and demographic features are less important than biomedical observations in the acute setting. After validation in other health system populations, our model could be implemented in clinical practice to enable better identification of patients with severe COVID-19 outcome.

## Methods

### Study design and setting

This retrospective study analyzed data gathered from Providence St. Joseph Health (PSJH), a community health system with 51 hospitals and 1085 clinics across five states in the western United States: Alaska, California, Montana, Oregon, and Washington. Inclusion criteria was age ≥18 years and confirmation of COVID-19 by a positive PCR-based SARS-CoV-2 test result. This study was performed in compliance with the Health Insurance Portability and Accountability Act (HIPAA) Privacy Rule and was approved by the Institutional Review Board (IRB) at PSJH with Study Number STUDY2020000196 with waiver of consent. We follow STROBE reporting guidelines (Supplemental Table 5).

### Task Definition

In this study, we hypothesized that age-stratified risk models for hospitalized patients with COVID-19 can accurately predict critical illness and mortality due to COVID-19 based on readily available patient data. Outcomes of patients were defined using the World Health Organization Ordinal Scale (WOS), proposed by the WHO R&D Blueprint group in their COVID-19 Therapeutic Trial Synopsis.^19^ The WHO ordinal scale ranges from 0 (uninfected) to 8 (deceased) with gradations depending on hospitalization, supplemental oxygen, mechanical ventilation, and organ support (vasopressors, renal replacement therapy, and extracorporeal membrane oxygenation). See Supplemental Table 1. In this study, we categorized WHO ordinal scores of 3-5 as the mild cases of COVID-19 and WHO ordinal scores of 6-8 as the critical illness and death within hospitalized patients. The objective is to develop machine learning models to predict critical illness and death with COVID-19 in hospitalized patients using easily available variables, including aggregated laboratory biomarkers and vital signs within one hour of either admission to the hospital or the first positive inpatient SARS-CoV-2 test. These predictive models are developed and tested on time horizons for one, seven, 14, 28, and 56 days from the confirmation of the infection and hospitalization. We compare the performance of machine learning models for 1) all-ages population, and 2) age-stratified subpopulations, to report the effect of age and compare the relative importance of risk factors between younger and aging adults.

### Population

The start time point of study is defined as June 31, 2021, after the delta became the predominant SARS-CoV-2 variant in the Western United States. Studied population included hospitalized individuals who received a positive test for COVID-19 between June 31, 2021 and November 15, 2021. This was confirmed by reverse-transcriptase polymerase chain reaction (RT-PCR) for the SARS-CoV-2 ribonucleic acid (RNA). Patients were excluded if they were already receiving mechanical ventilation at the time of admission to the hospital.

### Variables

The factors analyzed for prediction of COVID-19 outcomes were demographic characteristics, medical history, vital signs, and laboratory biomarkers (n = 64). Medical conditions included known risk factors for poor COVID-19 outcomes reported in the literature^5^ and conditions which are prevalent in aging patients (Table 1). Comorbidities that are usually chronic, such as hypertension, were included if they were active at the time of admission. Other comorbidities were included if they had been active any time within 2 years prior to admission, except for malignancy, which was included if active any time within the past 5 years. Note that active conditions mean health issues that affect the individual’s current functioning and all health. For biomedical precision, we used the Systematized Nomenclature of Medicine Clinical Terms (SNOMED–CT©) hierarchy (Supplemental Table 2), which can be mapped to ICD-10. Laboratory results and vital signs were included (both inpatient and outpatient) if they were collected between 24 hours before and one hour after either admission to the hospital or the first positive inpatient SARS-CoV-2 test (Table 1). Note that, we used aggregated temporal longitudinal vital signs in our model as described in Lee, et.al.^20^ Additionally, the risk factor list included patients need for supplemental oxygen mode, need for vasopressors, total number of comorbidities, and COVID-19 vaccination status.

**Table 1.** Demographics, vital signs, laboratory tests, and medical conditions analyzed for SARS-CoV-2 positive patients.

| Demographics | Vital signs | Laboratory tests | Medical conditions | Other Risk factors |
| --- | --- | --- | --- | --- |
| Age<br>Body mass index (BMI)<br>Sex<br>Reported ethnicity<br>Reported race | Heart rate (HR)<br>Respiratory rate (RR)<br>Systolic blood pressure (SBP)<br>Diastolic blood pressure (DBP)<br>Temperature<br>Oxygen saturation (SpO<sub>2</sub>) | White blood cell count (WBC)<br>Platelets (PLT)<br>Hematocrit (HCT)<br>Hemoglobin (HGT)<br>Basophils (BASO)<br>Eosinophils (EOSABS)<br>Lymphocytes (LYMABS)<br>Monocytes (MONO)<br>Neutrophils (NEUABS)<br>Potassium (K)<br>Sodium (NA)<br>Chloride (Cl)<br>Bicarbonate (HCO<sub>3</sub>)<br>Creatinine (CREA)<br>Blood urea nitrogen (BUN)<br>Glucose (GLU)<br>Albumin (ALB)<br>Alkaline (ALP)<br>Aspartate aminotransferase (AST)<br>Alanine aminotransferase (ALT)<br>Anion Gap (AGAP)<br>Bilirubin (TBIL)<br>Calcium (CA)<br>Globulin (GLOB)<br>Total Protein<br>D-dimer<br>C-reactive protein<br>Prothrombin time<br>BUN/Creatinine Ratio<br>Ferritin<br>International normalized ratio (INR)<br>Magnesium (MG)<br>Procalcitonin<br>Lactate dehydrogenase (LDH) | Hypertension<br>Coronary arteriosclerosis<br>Heart failure<br>Cardiomyopathy<br>Chronic obstructive pulmonary disease (COPD)<br>Asthma<br>Malignancy<br>Liver disease<br>Hyperlipidemia and Dyslipidemia<br>Obstructive sleep apnea<br>Chronic kidney disease<br>Diabetes mellitus<br>Solid organ transplant<br>Conditions related to reduced immune response<br>Dementia (All Causes) | Initial oxygen mode<br>Total Number of comorbidities<br>Vaccination status<br>Vasopressors |

### Statistical Analysis

Descriptive analyses are presented as frequencies and percentage for categorical variables, and as mean and standard deviation (std) for numerical variables. Fisher exact test was applied to compare distributions of categorical variables. The differences between distributions of numerical variables were calculated using Mann Whitney U-test. Results were considered statistically significant at a (2-tailed) p-value < 0.1. All statistical analyses were completed using PySpark version 2.4.5.

### Risk Model Development

In data preprocessing for development of each risk model, we removed features with missing values greater than 20% (Supplemental Table 3). We used IterativeImputer from Scikit Learn version 0.24.0 for imputing missing data in numerical features.^21^ Missing values for comorbidities were assumed to be absent from the patient’s medical history and imputed with a constant number of 0. Outliers were detected by calculating the modified z-score based on median absolute deviation with a threshold of 3.5 and then these outliers were imputed by the median.

To build machine learning models, we randomly split the dataset into 80% training data and 20% testing data and analyzed each patient using multiple algorithms including logistic regression (LR), random forest classification (RF), Adaptive Boosting (AdaBoost), and Gradient Boosting Decision Tree (GBDT). The parameters for each model were optimized using a 10-fold cross-validation on the training set with the maximum scoring value for the area under receiver operating characteristic curve score (AUROC). We then balanced true and false positive rates by optimizing the probability threshold for each class.

To address collinearity between predictors, we compared the optimum performance of logistic regression using the least absolute shrinkage and selection operator (LASSO) feature selection method. For non-linear tree-based models all features were included. Performance of models was reported as the area under the receiver operating characteristic curve (AUROC), area under the precision-recall curve (AUPRC), true positive rate (TPR), true negative rate (TNR), predictive positive value (PPV), and negative predictive value (NPV). We reported the 95% confidence interval for performance metrics of the models using Wilcoxon statistics,^22^ and binomial interval^23^ for the area under the ROC and precision-recall curves, respectively. All ML models were applied using Spark version 2.4.5, in the Python interface. We presented the interpretation of the model with the highest relative performance, gradient boosting, using the Shapley additive explanations (SHAP) algorithm, which uses cooperative game theory to calculate the marginal contribution of each feature, and examines the feature influence on model prediction.^24^ Predictive models were reported following TRIPOD guidelines.^25^

## Results

### Baseline characteristics

In the Providence St Joseph cohort (described in Methods above), 6,906 patients with positive tests for SARS-CoV-2 were analyzed (Supplemental Figure 1). Percent female was 44·25 and mean age was 59·90 years (SD ± 17·83 years), with a range 18 to 90+ years old. The distribution of relative frequency of hospitalizations by age is shown in Supplemental Figure 1. We divided the patients into two age subgroups: younger (age ≥ 18 and < 50 years with 1,963 patients) and older (≥ 50 years with 4,943 patients). The reported variables for prognosis of COVID-19 critical illness are presented in Table 2, Supplemental Table 3, and Supplemental Table 4. For patients with age ≥ 18 and < 50 years, the variables that had statistically significant correlation with critical illness and death in patients with COVID-19 were BMI, age, heart failure, and cardiomyopathy. For patients with age ≥ 50 years, the statistically significant variables were BMI, age, sex, dementia, and use of vasopressors within one hour of either admission to the hospital with COVID-19 or a first positive inpatient SARS-CoV-2 test. Vital signs values were aggregated from 24 hours before to one hour after and included (mean and standard deviation) for heart rate (HR), systolic blood pressure (SBP), diastolic blood pressure DBP, respiratory rate (RR), blood oxygen saturation (SpO2), and body temperature.

**Table 2:**
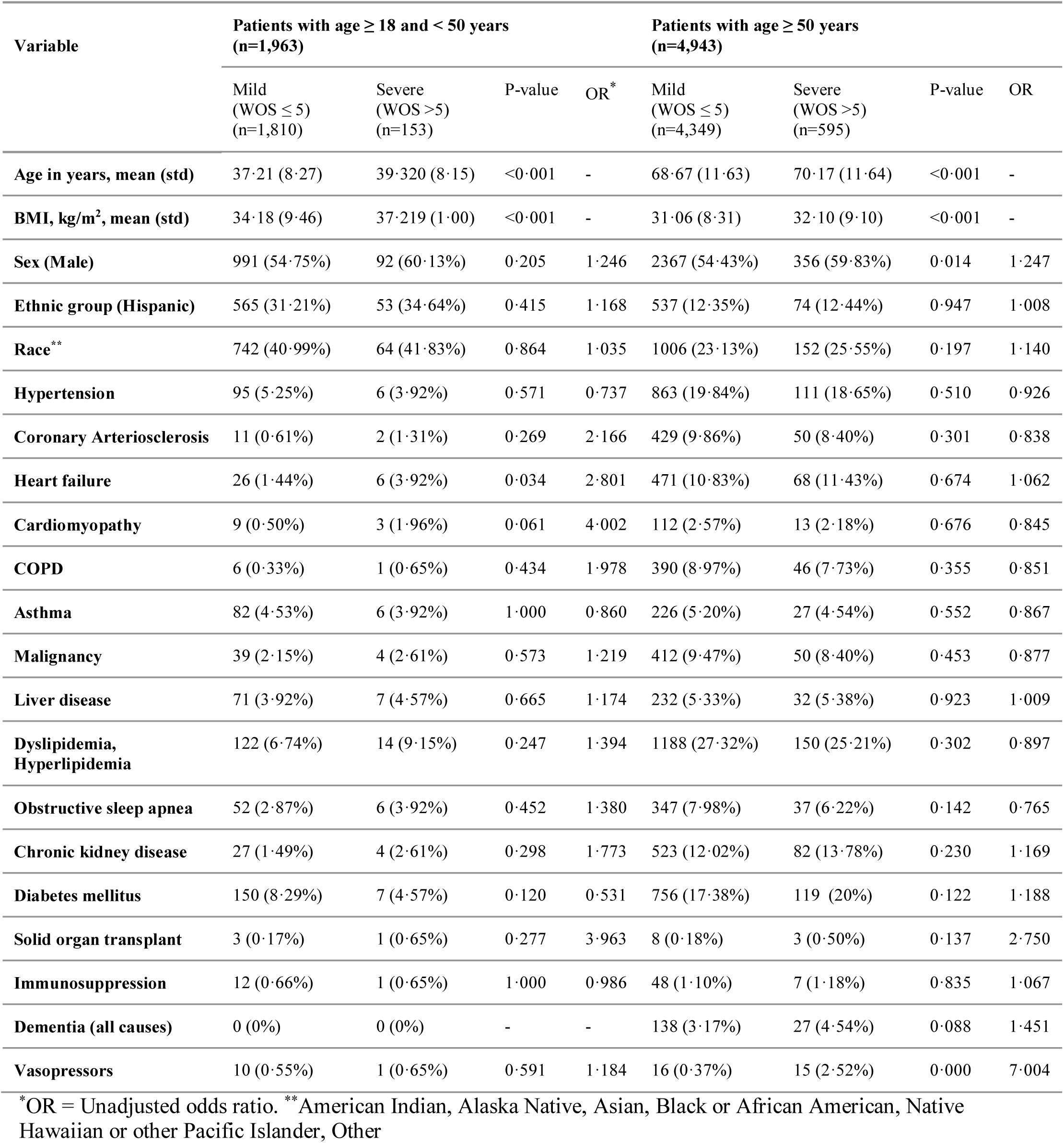
Demographics and medical conditions among hospitalized patients with COVID-19 by severity.

| Variable | Patients with age $\geq 18$ and $< 50$ years<br>(n=1,963) | | | | Patients with age $\geq 50$ years<br>(n=4,943) | | | |
| --- | --- | --- | --- | --- | --- | --- | --- | --- |
| | Mild<br>(WOS $\leq 5$)<br>(n=1,810) | Severe<br>(WOS $>5$)<br>(n=153) | P-value | OR* | Mild<br>(WOS $\leq 5$)<br>(n=4,349) | Severe<br>(WOS $>5$)<br>(n=595) | P-value | OR |
| Age in years, mean (std) | 37.21 (8.27) | 39.320 (8.15) | $<0.001$ | - | 68.67 (11.63) | 70.17 (11.64) | $<0.001$ | - |
| BMI, kg/m<sup>2</sup>, mean (std) | 34.18 (9.46) | 37.219 (1.00) | $<0.001$ | - | 31.06 (8.31) | 32.10 (9.10) | $<0.001$ | - |
| Sex (Male) | 991 (54.75%) | 92 (60.13%) | 0.205 | 1.246 | 2367 (54.43%) | 356 (59.83%) | 0.014 | 1.247 |
| Ethnic group (Hispanic) | 565 (31.21%) | 53 (34.64%) | 0.415 | 1.168 | 537 (12.35%) | 74 (12.44%) | 0.947 | 1.008 |
| Race** | 742 (40.99%) | 64 (41.83%) | 0.864 | 1.035 | 1006 (23.13%) | 152 (25.55%) | 0.197 | 1.140 |
| Hypertension | 95 (5.25%) | 6 (3.92%) | 0.571 | 0.737 | 863 (19.84%) | 111 (18.65%) | 0.510 | 0.926 |
| Coronary Arteriosclerosis | 11 (0.61%) | 2 (1.31%) | 0.269 | 2.166 | 429 (9.86%) | 50 (8.40%) | 0.301 | 0.838 |
| Heart failure | 26 (1.44%) | 6 (3.92%) | 0.034 | 2.801 | 471 (10.83%) | 68 (11.43%) | 0.674 | 1.062 |
| Cardiomyopathy | 9 (0.50%) | 3 (1.96%) | 0.061 | 4.002 | 112 (2.57%) | 13 (2.18%) | 0.676 | 0.845 |
| COPD | 6 (0.33%) | 1 (0.65%) | 0.434 | 1.978 | 390 (8.97%) | 46 (7.73%) | 0.355 | 0.851 |
| Asthma | 82 (4.53%) | 6 (3.92%) | 1.000 | 0.860 | 226 (5.20%) | 27 (4.54%) | 0.552 | 0.867 |
| Malignancy | 39 (2.15%) | 4 (2.61%) | 0.573 | 1.219 | 412 (9.47%) | 50 (8.40%) | 0.453 | 0.877 |
| Liver disease | 71 (3.92%) | 7 (4.57%) | 0.665 | 1.174 | 232 (5.33%) | 32 (5.38%) | 0.923 | 1.009 |
| Dyslipidemia,<br>Hyperlipidemia | 122 (6.74%) | 14 (9.15%) | 0.247 | 1.394 | 1188 (27.32%) | 150 (25.21%) | 0.302 | 0.897 |
| Obstructive sleep apnea | 52 (2.87%) | 6 (3.92%) | 0.452 | 1.380 | 347 (7.98%) | 37 (6.22%) | 0.142 | 0.765 |
| Chronic kidney disease | 27 (1.49%) | 4 (2.61%) | 0.298 | 1.773 | 523 (12.02%) | 82 (13.78%) | 0.230 | 1.169 |
| Diabetes mellitus | 150 (8.29%) | 7 (4.57%) | 0.120 | 0.531 | 756 (17.38%) | 119 (20%) | 0.122 | 1.188 |
| Solid organ transplant | 3 (0.17%) | 1 (0.65%) | 0.277 | 3.963 | 8 (0.18%) | 3 (0.50%) | 0.137 | 2.750 |
| Immunosuppression | 12 (0.66%) | 1 (0.65%) | 1.000 | 0.986 | 48 (1.10%) | 7 (1.18%) | 0.835 | 1.067 |
| Dementia (all causes) | 0 (0%) | 0 (0%) | - | - | 138 (3.17%) | 27 (4.54%) | 0.088 | 1.451 |
| Vasopressors | 10 (0.55%) | 1 (0.65%) | 0.591 | 1.184 | 16 (0.37%) | 15 (2.52%) | 0.000 | 7.004 |

\*OR = Unadjusted odds ratio. \*\*American Indian, Alaska Native, Asian, Black or African American, Native Hawaiian or other Pacific Islander, Other

### Risk Model Analysis

In this paper, we trained five ML models including LR, RF, GBDT, and AdaBoost for the all-age population (n=6,906), and two different age subpopulations (patients with age ≥ 18 and < 50 years with n=1,963 and patients with age ≥ 50 years with n=4,943) using the aggregated values of predictors. Class distribution for outcomes show that patients with critical illness and death accounted for 7·79% of the younger cohort with age ≥ 18 and < 50 years and 12·04% of the older cohort with age ≥ 50 years. This class imbalance was addressed by undersampling patients with mild severity from the training set. Results were reported on the complete test dataset, representing actual population distribution. Supplemental Table 5 represents the performance results for three sets of developed models for younger, older patients and all-age groups. These performance results were reported after adjusting the probability threshold to optimize models for clinical and research applications. Among four models for the younger population, GBDT had the highest true positive rate of 67·73%, true negative rate of 67·40%, and AUROC value of 0·80. For the older population, GBDT had a maximum true positive rate of 76·92%, true negative rate of 76·95% and AUROC of 0·83. Figure 1 represents the comparison between the AUROC value for four ML models based on the patient’s age. Relative feature importance for the younger, older and generalized GBDT models was determined by Shapley additive explanations (SHAP), as shown in Figure 2, and Supplemental Figure 3, respectively. SHAP values were also used to assess the contribution of age on each model outcome (Supplemental Figure 3). We used the distribution of importance for each variable to assess its contribution to model outcome. In the younger population, some variables for comorbidities added no predictive value, which resulted in them being automatically removed from the SHAP plot.

**Figure 1:**
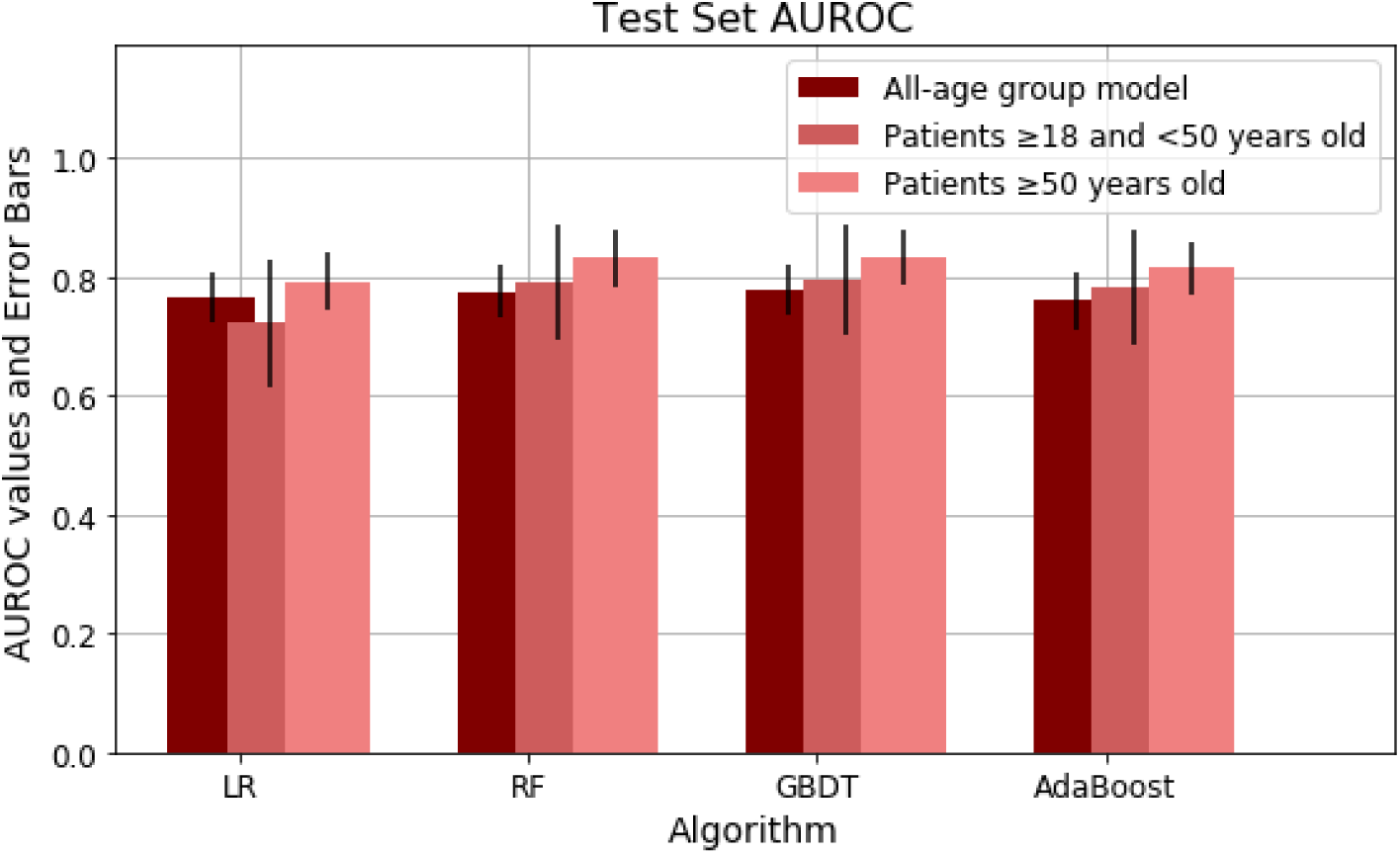
Area under receiver operator characteristic curve (AUROC) for age-stratified models of severe COVID-19 outcomes in hospitalized patients.

**Figure 2:**
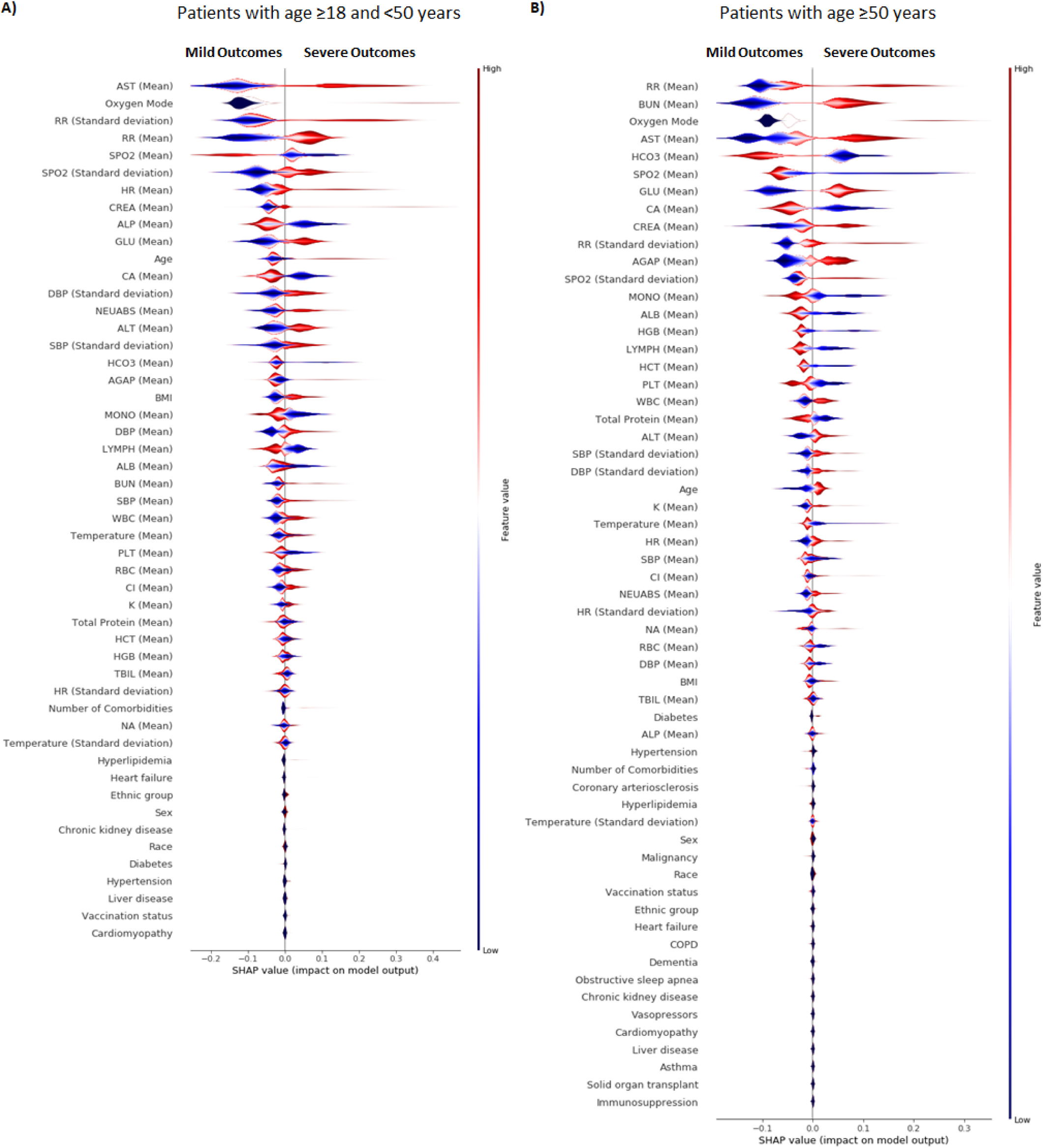
Gradient Boosting Decision Tree feature importance for age-stratified models of severe COVID-19 outcomes in hospitalized patients. A) Feature importance and the influence of higher and lower values of the risk factors on the patient with age ≥ 18 and < 50 years outcome, B) Feature importance and the influence of higher and lower values of the risk factors on the patient with age ≥ 50 years outcome. Note that the left side of this graph represents reduced risk of critical illness or death, and the right side of the graph represents the increased risk of critical illness and death outcome. Nominal classes are binary [0,1]. For sex, female is 0 (blue) and for race, White is 0 (blue).

Additionally, we used the GBDT to validate and assess the performance of the model for different time horizons. For the all-age group, gradient boosting showed an AUROC value of 0·80, 0·78, 0·77 and 0·77 for respectively, one, 14, 28 and 56 day intervals after the confirmation of infection. Furthermore, we predicted the mortality of patients (WHO ordinal score of 8) using the GBDT model and the full set of aggregated risk factors. Note that to predict the mortality of patients with COVID-19, we also included the patients who were already receiving mechanical ventilation and additional organ support (WHO ordinal score of 6 and 7), see Supplemental Figure 1. Therefore, the number of all age group patients for predicting mortality increased to 7,063. The results show the AUROC value of 0·85 for the general population and 0·76 and 0·80 for the younger and aging population, respectively.

## Discussion

In this study, we developed risk models to predict the outcomes of hospitalized adult patients with COVID-19, in the context of current COVID-19 standard of care and delta variant predominance. We used clinical data from within one hour of either admission to the hospital with COVID-19 or the first positive inpatient SARS-CoV-2 test result. Explainability analysis on the machine learning models showed that risk factors are different for older patients compared to younger patients. This is the first study that investigates age-stratified modeling for COVID-19 severity for hospitalized adults for early prediction across multiple time horizons. Data from 6,906 patients across five states was used to develop predictive models for COVID-19 critical illness and death in younger and older hospitalized adults within one, seven, 14, 28 and 56 days of positive infection test and hospitalization. The key findings are: 1) risk models perform well using readily available clinical data, 2) vital signs and laboratory results at the time of admission are more important for prediction than the presence of comorbidities, 3) age-stratified models show that the relative importance of risk factor differs between younger and older adults.

Since the beginning of the pandemic, standard of COVID-19 care has improved and delta has become the predominant variant. Further, risk models from earlier in the pandemic relied on labs that are not routinely used in many patients.^26^ This was reflected by the high rate of missing values for tests required for in early risk scores, including INR, D-dimer, ferritin and procalcitonin (PCT). The models developed here are both performant and pragmatic.

Our statistical analysis revealed new insights on how variables that correlate significantly with critical illness and death in COVID-19 differ between younger and older age groups. For example, most comorbidities such as malignancy, cardiomyopathy and COPD have higher odds ratios for severe outcomes in younger patients than in older patients. Conversely, lower BUN/creatinine ratio and lower potassium are only statistically associated with critical illness and death in older patients.

We chose GBDT, a sequential ensemble approach,^27^ as the model with the best relative performance to define the most predictive variables for COVID-19 outcomes. Non-linear models showed higher performance than linear models, suggesting better representation of complex interactions across multiple mechanisms of disease. Stratifying patients by age group revealed that, in general, vital signs and laboratory tests have a higher relative importance than comorbidities. Because age is such a significant risk factor, it can mask other important predictors. By removing the confounding effects of age, these models highlight new insights into risk factors for IMV and death.

Additionally, we investigated the effect of age on predictive models for younger and older COVID-19 patients. For patients with age ≥ 18 and < 50 years (Supplemental Figure 3C), age has a relatively high and more consistent predictive effect on the performance of the model. Within patients younger than 50 years old, higher age had a negative effect on outcome. However, in patients with age ≥ 50 years (Supplemental Figure 3D), age has less effect on the model performance. Patient stratification removed some of the confounding effect of age^28^ in this group, better revealing the contribution of laboratory results, vital signs and comorbidities as predictors.

For the younger population, the patient’s initial oxygen mode and aggregated vital signs demonstrate the highest predictive value for outcome severity. Other predictive factors include higher AST, higher creatinine, lower alkaline phosphatase, and lower calcium levels, higher age, and higher BMI. Laboratory results have higher importance for older patients than they do for younger patients. Features such as higher BUN, higher AST, lower HCO3, lower calcium, and some aggregated vital signs (respiratory rate, blood pressure and SpO2) are among the most predictive. Sex is not a strong predictive factor, despite it having an odds ratio of ∼1·25 in both the older and younger population. BMI is another feature that supports the importance of analyzing age subgroups separately. It is statistically correlated to the severity of COVID-19 and is an important predictor for the younger population but shows no significant correlation in the older population (Supplemental Figure 3). This could be explained by higher BMI in younger hospitalized patients compared to the older hospitalized patients with COVID-19.^29^ Future investigation is needed to determine risks with being underweight or overweight, potentially with BMI-stratified models. Neither race or ethnicity had strong feature importance for prediction in the younger and older population. This shows that although chronic comorbidities (binary diagnostic labels), BMI, sex, race, ethnicity may have high odds ratios in a univariate analysis, these factors are much less important in the acute setting for predicting critical illness. Once hospitalized, biomedical observations are more predictive.

SHAP values also indicate the direction of variables’ impact on outcomes. For example, higher serum creatinine levels, lower platelet counts, lower lymphocyte counts, and higher neutrophil count are all predictive of critical illness and death.^33–35^ Lower calcium is associated with more severe COVID-19, as noted in previous studies,^30^ and this analysis shows it has higher predictive value in older patients.

Hence, age stratification shows that risk factors for severe COVID-19 differ by age, in ways that cannot be determined in all-age models. This affirms the importance of analyzing each different age group separately, particularly for the older population who have the greater overall risk for poor outcomes.

Also, as expected, vaccination reduced the risk of severe outcomes in the older population. Vaccination status had relatively low importance, which may reflect the low number of hospitalized patients who had received vaccination during the observation window; only 8·10% of the younger hospitalized patients and 25·48% of older hospitalized patients had received at least one dose of a vaccine.

Early risk stratification in patients with COVID-19 is essential to inform decisions about what level of care a patient is likely to need. One of the main challenges of COVID-19 is the heterogeneity of presentation; therefore factors related to poor outcomes are not always evident at admission.^13^ In this study, ML models using readily available variables (demographics, vital signs, common laboratory test and medical history) demonstrated strong performance for predicting the severity of COVID-19. Importantly, the population in this study included patients from 51 hospitals and 1081 clinics across five states, using data based on the current standard of care for COVID-19 and the delta variant. Five limitations of this retrospective study are: 1) reliance on EHR structured data which can miss medical conditions that not diagnosed, not recorded, or noted only in free text, 2) use of hospital reported race and ethnicity of patients^31^ as opposed to direct per-patient measures of potential confounders (genetic information, disparities in healthcare, and individual lifetime history of beneficial and harmful exposures, 4) use of data from within a single healthcare system. Concerns regarding generalizability of this study are partially mitigated by the size and diversity of PSJH, which serves both urban and rural communities from California to Alaska. Future investigations will benefit from finer granularity of subdivisions by age, BMI, and more detailed variables on conditions and drugs that affect individual immune response.

## Conclusion

We developed two age-stratified risk models for critical illness in hospitalized patients with COVID-19 and tested them on data from patients during times of improved standard of care treatment and delta variant predominance. For hospitalized adults, baseline data that is readily available within one hour after hospital admission or a first positive inpatient SARS-CoV-2 test can predict critical illness within one day, and up to 56 days later. The models for age ≥ 18 and < 50 years and the model for age ≥ 50 years were both more performant than all-age models. These age-stratified models also revealed differences in the statistical significance and relative predictive value of risk factors between older and younger patients, including age, BMI, vital signs, and laboratory results. In addition, sex and chronic comorbidities had lower predictive value than vital signs and laboratory results. The results of this age-stratified modeling approach provide advanced understanding of current risk factors for severe COVID-19 outcomes and can help inform care decisions and prioritize next steps for research.

## Data Availability

All data produced in the present study are available upon reasonable request to the authors.

## Conflict of Interest Disclosures

None of the authors have a conflict of interest with this study.

## Funding/Support

This work was funded by NIH NIA grant 2U01AG046139-06 (to NDP, NET, and TEG). JJH and VRD have been funded in part with Federal funds from the Department of Health and Human Services, Office of the Assistant Secretary for Preparedness and Response, Biomedical Advanced Research and Development Authority, under Contract No. HHSO100201600031C, administered by Merck, Inc., on work unrelated to this study. SM and JJH have been funded through Pfizer Inc. Grant No. 68114891 on work unrelated to this study. NET is also funded by NIH NIA R01 AG061796.

## Role of the Funder/Sponsor

The funders had no role in the design and conduct of the study; collection, management, analysis, and interpretation of the data; preparation, review, or approval of the manuscript; and decision to submit the manuscript for publication.

## Acknowledgements/Contributions

SM, PVH, JDG, and JJH conceptualized the study. SM, RTR, PVH, VRD and AMB were involved in the EHR data extraction, data cleaning, and codification. SM performed data analysis including statistical analysis, machine learning and data interpretation. JJH supervised implementation. Administrative and material support was provided by JJH and NDP. SM and PVH prepared the manuscript with critical revision of the manuscript for important intellectual content provided by JDG, NR, and JJH. All authors reviewed and approved the final version of the manuscript. We would also like to acknowledge SNOMED International for developing and maintaining SNOMED CT.

---

**Supplemental Figure 1:**
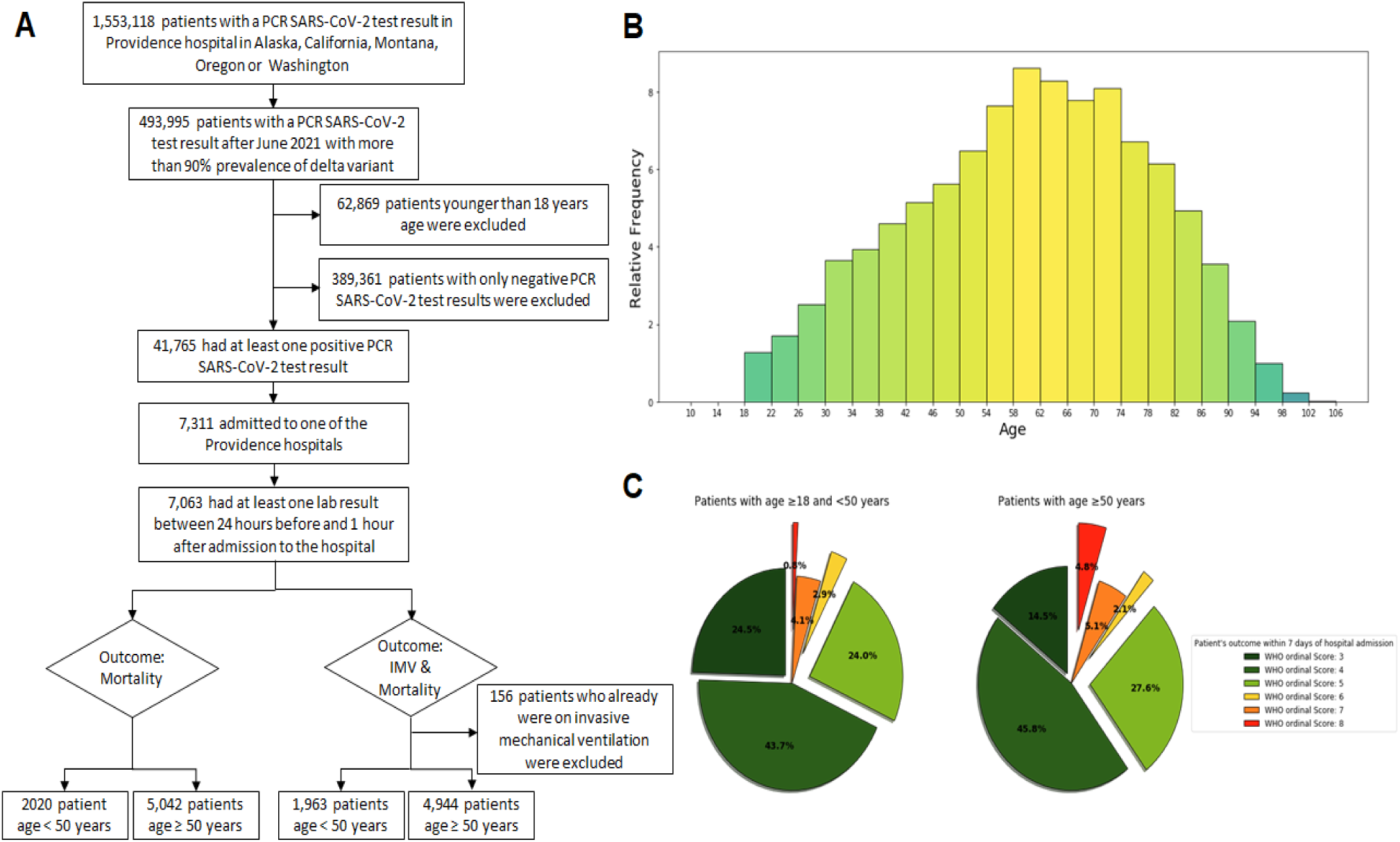
Study cohort. (A) Cohort selection from PSJH-EHR data. (B) The frequency histogram of hospitalized patients based on their age. (C) The frequency of hospitalized patients maximum WHO ordinal score within 7 days of hospitalization based on their age.

**Supplemental Figure 2:**
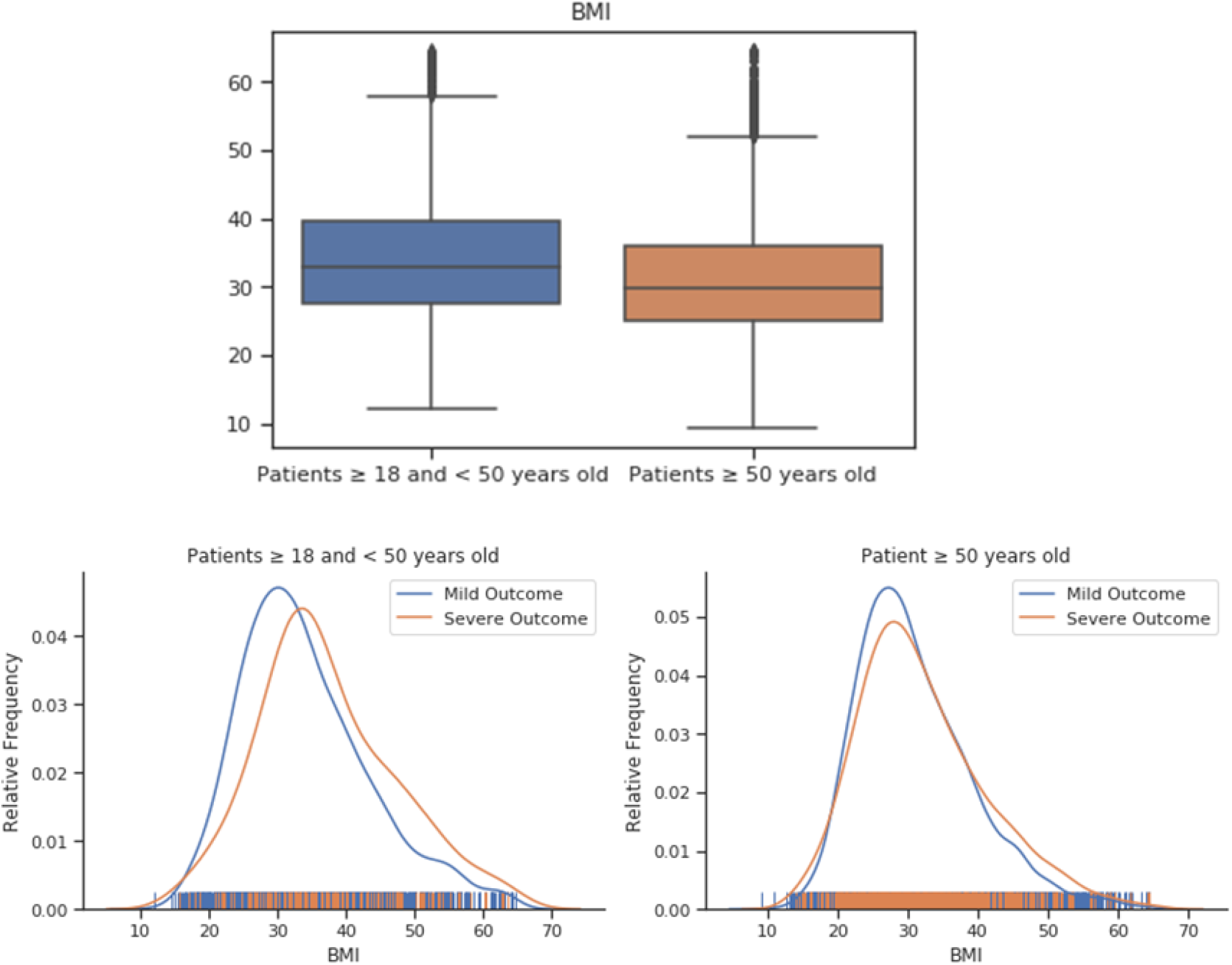
Distribution of patients’ BMI for invasive mechanical ventilation and mortality outcome in different age groups.

**Supplemental Figure 3:**
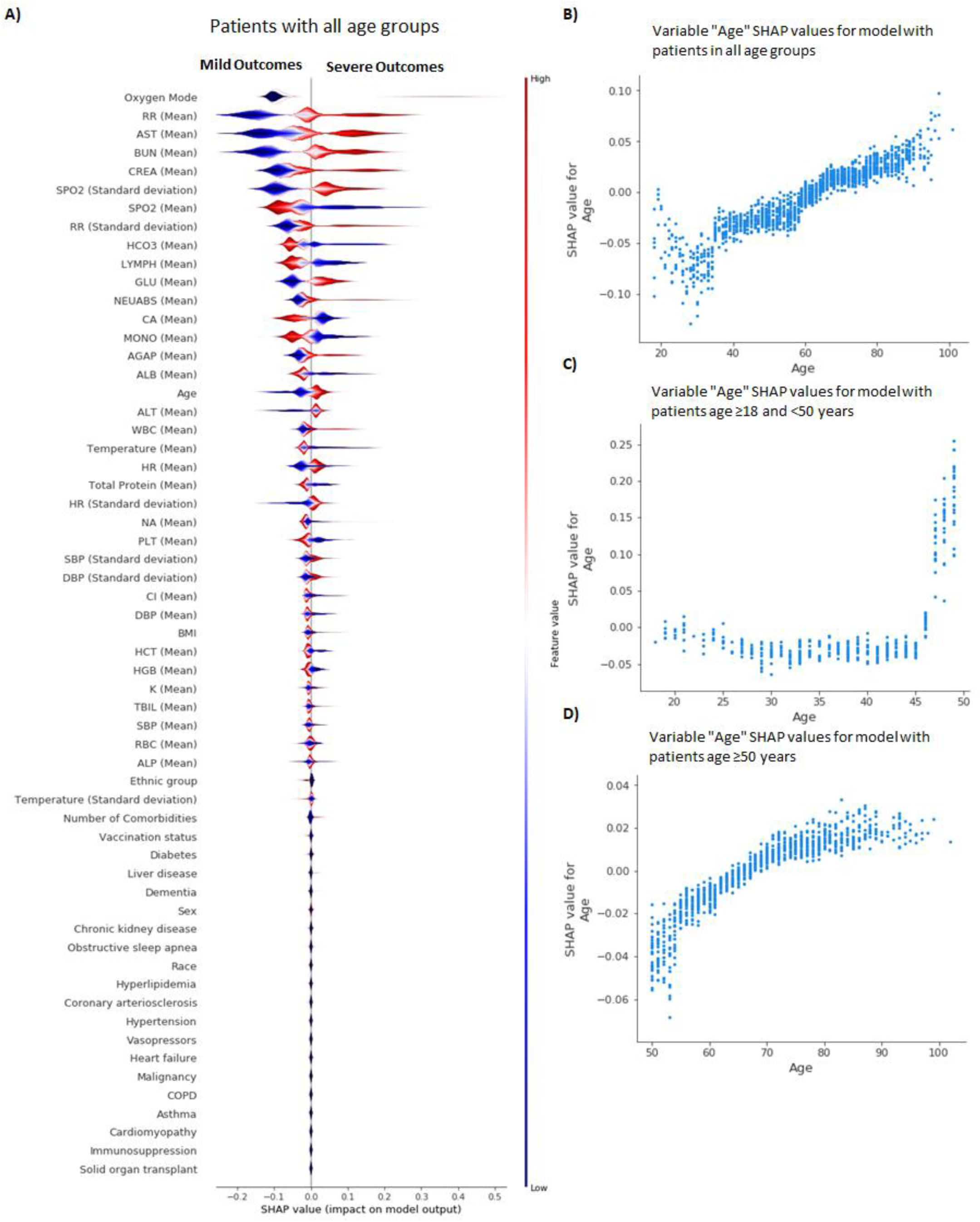
Feature importance for all-age model of severe COVID-19 outcomes in hospitalized patients. A) Gradient Boosting Decision Tree feature importance and the influence of higher and lower values of the risk factors on the all-age group population outcome. Note that the left side of this graph represents reduced risk of critical illness and death outcome and right side of the graph represents the increased risk of critical illness and death outcome. Nominal classes are binary [0,1]. For sex, female is 0 (blue) and for race, White is 0 (blue), B: Variable “Age” contribution in prediction process of model for patients’ with all age groups, C: Variable “Age” contribution in prediction process of model for patients’ age ≥ 18 and < 50 years, D: Variable “Age” contribution in prediction process of model for patients’ age ≥ 50 years.

**Supplemental Table 1:** Implementation of World Health Organization Ordinal Scale (WOS)

| Patient State | Descriptor | Score | Implementation |
| --- | --- | --- | --- |
| **Uninfected** | No clinical or virological evidence | 0 | Not applicable: all patients in study were hospitalized. |
| **Ambulatory** | No limitation of activities | 1 | Not applicable: all patients in study were hospitalized. |
|  | Limitation of activities | 2 | Not applicable: all patients in study were hospitalized. |
| **Hospitalized mild disease** | Hospitalized, no oxygen therapy | 3 | Encounter |
|  | Oxygen by mask or nasal prongs | 4 | Nursing flowsheets, step function |
|  | Non-invasive ventilation of high-flow oxygen | 5 | Nursing flowsheets, step function |
| **Hospitalized severe disease** | Intubation and mechanical ventilation | 6 | Nursing flowsheets, step function |
|  | Ventilation and additional organ support: pressors, renal replacement therapy, dialysis, extracorporeal membrane oxygenation (ECMO) | 7 | Vasopressor in medication administration records. RxNorm ingredient: 3616 dobutamine, 3628 dopamine, 3966 ephedrine, 3992 epinephrine, 6963 midodrine, 7512 norepinephrine, 8163 phenylephrine, 11149 vasopressin (USP)<br>Renal replacement therapy: Nursing flowsheets, step function<br>ECMO: Nursing flowsheets, step function |
|  | Death | 8 | Date of death is recorded |

**Supplemental Table 2:**
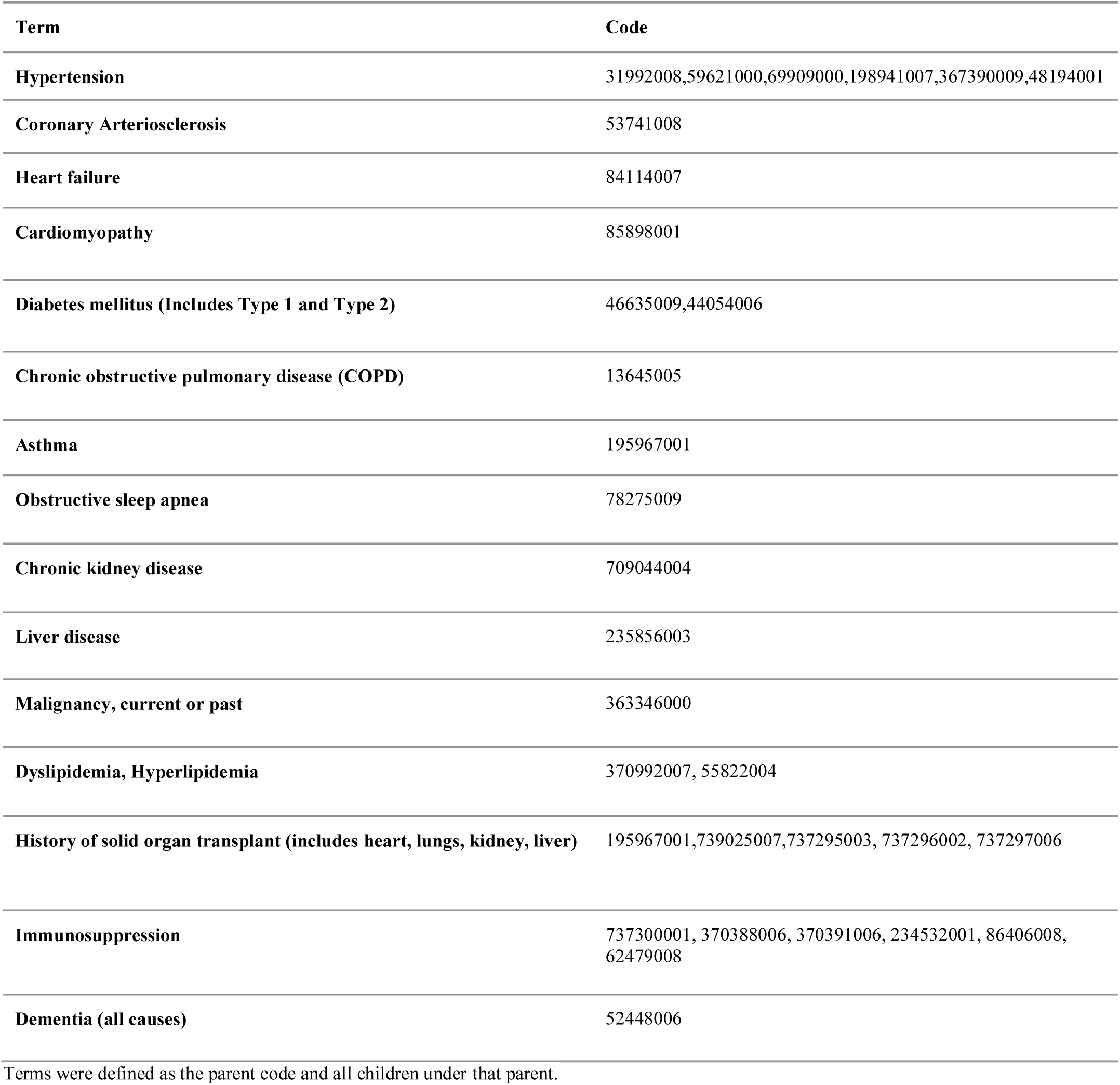
Terms and SNOMED-CT codes.

**Supplemental Table 3:**
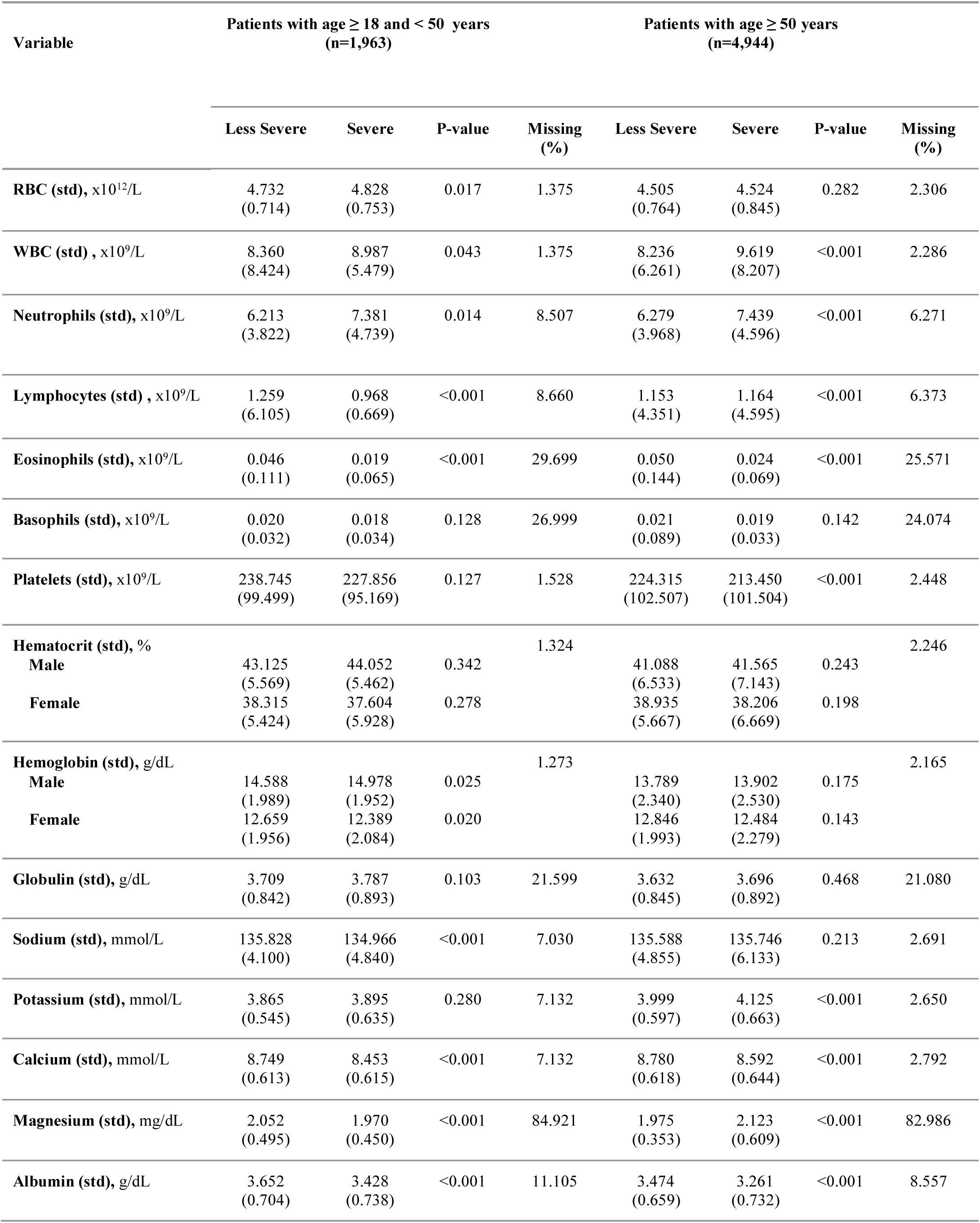
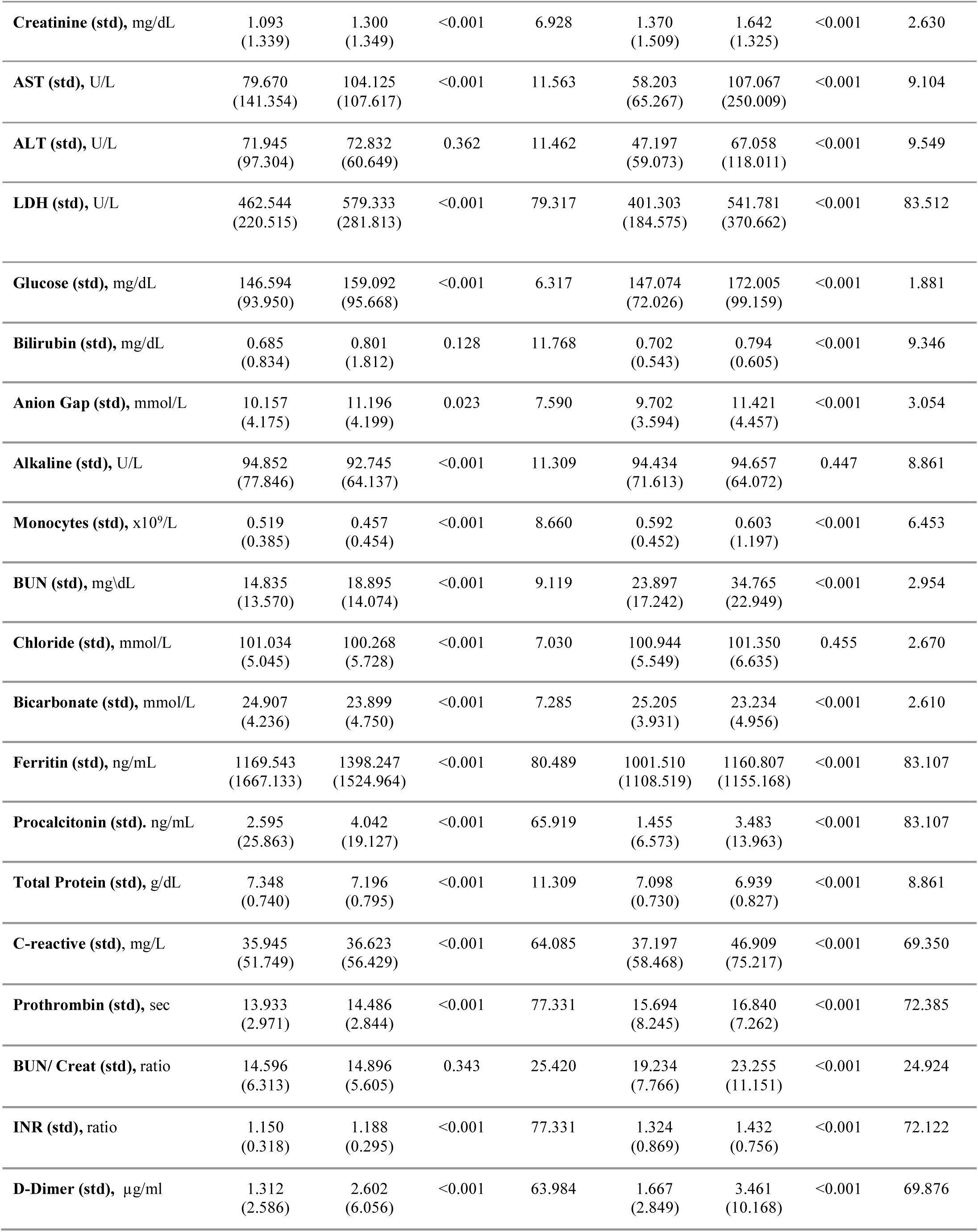
Mean and standard deviation values and missing percentage of laboratory results among hospital patients with COVID-19 by severity.

**Supplemental Table 4:** Vital sign data among patients with COVID-19 according to the severity.

| Variable | Patients $\geq 18$ and $< 50$ years<br>(n=706) | | | Patients $\geq 50$ years old<br>(n=2531) | | |
| --- | --- | --- | --- | --- | --- | --- |
|  | Less Severe | Severe | P-value | Less Severe | Severe | P-value |
| **SBP** | 124.848 (17.381) | 125.949 (18.416) | 0.143 | 130.859 (20.502) | 128.440 (22.401) | <0.001 |
| **DBP** | 75.321 (11.774) | 76.437 (12.250) | 0.130 | 73.193 (12.219) | 71.963 (12.723) | <0.001 |
| **RR** | 22.422 (5.631) | 27.148 (8.235) | <0.001 | 21.747 (4.708) | 25.363 (5.960) | <0.001 |
| **SpO<sub>2</sub>** | 94.341 (3.441) | 91.086 (5.162) | <0.001 | 93.821 (3.408) | 91.751 (5.021) | <0.001 |
| **HR** | 97.374 (16.404) | 106.390 (16.577) | <0.001 | 88.989 (16.821) | 94.054 (19.447) | <0.001 |
| **Body Temp.** | 99.178 (1.652) | 99.432 (1.594) | 0.013 | 98.839 (1.678) | 98.863 (1.613) | 0.080 |

**Supplemental Table 5:** Prediction performance and 95% confidence interval on the test set for models with full set of features.

| Age group | Classifier | AUROC | PR-AUC | TPR | TNR | PPV | NPV | LKR+ | LKR- |
| --- | --- | --- | --- | --- | --- | --- | --- | --- | --- |
| **Generalized model for patients in all age groups** | LR | 0.7672<br>(0.72,0.81) | 0.4606<br>(0.38,0.54) | 0.6776<br>(0.60,0.75) | 0.6813<br>(0.65,0.71) | 0.2081<br>(0.17,0.24) | 0.9447<br>(0.93,0.96) | 2.1261<br>(1.85,2.44) | 0.4732<br>(0.37,0.60) |
|  | RF | 0.7764<br>(0.73,0.82) | 0.4876<br>(0.41,0.57) | 0.7105<br>(0.64,0.78) | 0.7106<br>(0.65,0.74) | 0.2328<br>(0.19,0.27) | 0.9521<br>(0.94,0.97) | 2.4551<br>(2.15,2.81) | 0.4074<br>(0.32,0.52) |
|  | GBDT | 0.7784<br>(0.73,0.82) | 0.4868<br>(0.41,0.57) | 0.7105<br>(0.64,0.78) | 0.7081<br>(0.68,0.73) | 0.2313<br>(0.19,0.27) | 0.9519<br>(0.94,0.97) | 2.4341<br>(2.13,2.78) | 0.4088<br>(0.32,0.53) |
|  | AdaBoost | 0.7603<br>(0.71,0.81) | 0.4702<br>(0.39,0.55) | 0.6908<br>(0.62,0.76) | 0.6894<br>(0.66,0.71) | 0.2156<br>(0.18,0.25) | 0.9475<br>(0.93,0.96) | 2.2241<br>(1.94,2.55) | 0.4485<br>(0.35,0.57) |
| **Age-stratified model for patients age $\geq 18$ and $< 50$ years** | LR | 0.7222<br>(0.62,0.83) | 0.3965<br>(0.22,0.57) | 0.6451<br>(0.48,0.81) | 0.5939<br>(0.54,0.64) | 0.1198<br>(0.07,0.17) | 0.9513<br>(0.92,0.98) | 1.5885<br>(1.19,2.12) | 0.5976<br>(0.37,0.97) |
|  | RF | 0.7930<br>(0.70,0.89) | 0.4238<br>(0.25,0.60) | 0.6773<br>(0.51,0.84) | 0.6574<br>(0.61,0.71) | 0.1449<br>(0.09,0.20) | 0.9596<br>(0.93,0.98) | 1.9769<br>(1.49,2.62) | 0.4909<br>(0.29,0.82) |
|  | GBDT | 0.7965<br>(0.70,0.89) | 0.4270<br>(0.25,0.60) | 0.6773<br>(0.51,0.84) | 0.6740<br>(0.63,0.72) | 0.1511<br>(0.09,0.21) | 0.9606<br>(0.94,0.98) | 2.0776<br>(1.56,2.76) | 0.4788<br>(0.29,0.80) |
|  | AdaBoost | 0.7840<br>(0.68,0.88) | 0.4085<br>(0.24,0.58) | 0.6451<br>(0.48,0.81) | 0.6713<br>(0.62,0.72) | 0.1439<br>(0.08,0.20) | 0.9567<br>(0.93,0.98) | 1.9626<br>(1.45,2.65) | 0.5287<br>(0.33,0.85) |
| **Age-stratified model for patients age $\geq 50$ years** | LR | 0.7920<br>(0.74,0.84) | 0.5210<br>(0.43,0.61) | 0.7350<br>(0.65,0.81) | 0.7408<br>(0.71,0.77) | 0.2757<br>(0.23,0.32) | 0.9542<br>(0.94,0.97) | 2.8356<br>(2.43,3.32) | 0.3577<br>(0.26,0.48) |
|  | RF | 0.8318<br>(0.79,0.88) | 0.5513<br>(0.46,0.64) | 0.7692<br>(0.69,0.85) | 0.7660<br>(0.74,0.79) | 0.3061<br>(0.25,0.36) | 0.9611<br>(0.95,0.97) | 3.2872<br>(2.81,3.84) | 0.3013<br>(0.22,0.42) |
|  | GBDT | 0.8336<br>(0.79,0.88) | 0.5529<br>(0.46,0.64) | 0.7692<br>(0.69,0.85) | 0.7695<br>(0.74,0.80) | 0.3093<br>(0.26,0.36) | 0.9613<br>(0.95,0.98) | 3.3371<br>(2.85,3.90) | 0.3999<br>(0.21,0.42) |
|  | AdaBoost | 0.8155<br>(0.77,0.86) | 0.5419<br>(0.45,0.63) | 0.7521<br>(0.67,0.83) | 0.7672<br>(0.74,0.79) | 0.3024<br>(0.25,0.35) | 0.9584<br>(0.94,0.97) | 3.2307<br>(2.75,3.79) | 0.3231<br>(0.23,0.44) |

AUROC = area under the receiver-operating characteristic curve. PR-AUC: area under the precision-recall curve. TPR = true positive rate. TNR = true negative rate. LKR+ = positive likelihood ratio. LKR- = negative likelihood ratio. PPV = positive predictive value. NPV = negative predictive value.

## Notes

### Competing Interest Statement

The authors have declared no competing interest.

### Author Declarations

Ethics IRB of Institute for Systems Biology gave ethical approval for this work.

## References

1. WHO Coronavirus Disease (COVID-19) Dashboard. Accessed October 8, 2020. https://covid19.who.int/

2. Bohn MK, Hall A, Sepiashvili L, Jung B, Steele S, Adeli K. Pathophysiology of COVID-19: Mechanisms Underlying Disease Severity and Progression. Physiology. 2020;35(5):288–301.

3. Joost Wiersinga W, Rhodes A, Cheng AC, Peacock SJ, Prescott HC. Pathophysiology, Transmission, Diagnosis, and Treatment of Coronavirus Disease 2019 (COVID-19): A Review. JAMA. 2020;324(8):782–793.

4. Zhou F, Yu T, Du R, et al. Clinical course and risk factors for mortality of adult inpatients with COVID-19 in Wuhan, China: a retrospective cohort study. Lancet. 2020;395(10229):1054–1062.

5. Gao Y-D, Ding M, Dong X, et al. Risk factors for severe and critically ill COVID-19 patients: A review. Allergy. 2021;76(2):428–455.

6. Farshbafnadi M, Kamali Zonouzi S, Sabahi M, Dolatshahi M, Aarabi MH. Aging & COVID-19 susceptibility, disease severity, and clinical outcomes: The role of entangled risk factors. Exp Gerontol. 2021;154:111507.

7. Bae S, Kim SR, Kim M-N, Shim WJ, Park S-M. Impact of cardiovascular disease and risk factors on fatal outcomes in patients with COVID-19 according to age: a systematic review and meta-analysis. Heart. 2021;107(5):373–380.

8. Burki TK. Lifting of COVID-19 restrictions in the UK and the Delta variant. Lancet Respir Med. 2021;9(8):e85.

9. Marcos M, Belhassen-García M, Sánchez-Puente A, et al. Development of a severity of disease score and classification model by machine learning for hospitalized COVID-19 patients. PLoS One. 2021;16(4):e0240200.

10. Lombardi Y, Azoyan L, Szychowiak P, et al. External validation of prognostic scores for COVID-19: a multicenter cohort study of patients hospitalized in Greater Paris University Hospitals. Intensive Care Med. Published online September 28, 2021. doi:10.1007/s00134-021-06524-w

11. King JT Jr, Yoon JS, Bredl ZM, et al. Accuracy of the Veterans Health Administration COVID-19 (VACO) Index for predicting short-term mortality among 1307 US academic medical centre inpatients and 427 224 US Medicare patients. J Epidemiol Community Health. Published online September 28, 2021. doi:10.1136/jech-2021-216697

12. Rinderknecht MD, Klopfenstein Y. Predicting critical state after COVID-19 diagnosis: model development using a large US electronic health record dataset. NPJ Digit Med. 2021;4(1):113.

13. Yadaw AS, Li Y-C, Bose S, Iyengar R, Bunyavanich S, Pandey G. Clinical features of COVID-19 mortality: development and validation of a clinical prediction model. Lancet Digit Health. 2020;2(10):e516–e525.

14. Nicholson CJ, Wooster L, Sigurslid HH, et al. Estimating risk of mechanical ventilation and in-hospital mortality among adult COVID-19 patients admitted to Mass General Brigham: The VICE and DICE scores. EClinicalMedicine. 2021;33:100765.

15. Knight SR, Ho A, Pius R, et al. Risk stratification of patients admitted to hospital with covid-19 using the ISARIC WHO Clinical Characterisation Protocol: development and validation of the 4C Mortality Score. BMJ. 2020;370. doi:10.1136/bmj.m3339

16. Chen Z, Chen J, Zhou J, et al. A risk score based on baseline risk factors for predicting mortality in COVID-19 patients. Curr Med Res Opin. 2021;37(6):917–927.

17. Garcia-Gordillo JA, Camiro-Zúñiga A, Aguilar-Soto M, et al. COVID-IRS: A novel predictive score for risk of invasive mechanical ventilation in patients with COVID-19. PLoS One. 2021;16(4):e0248357.

18. Ahmad MA, Eckert C, Teredesai A. Interpretable Machine Learning in Healthcare. In: Proceedings of the 2018 ACM International Conference on Bioinformatics, Computational Biology, and Health Informatics. BCB ‘18. Association for Computing Machinery; 2018:559–560.

19. Coronavirus N. WHO R&D Blueprint. https://www.who.int/blueprint/priority-diseases/key-action/COVID-19_Treatment_Trial_Design_Master_Protocol_synopsis_Final_18022020.pdf

20. Lee JY, Molani S, Fang C, et al. Ambulatory Risk Models for the Long-Term Prevention of Sepsis: Retrospective Study. JMIR Med Inform. 2021;9(7):e29986.

21. Pedregosa F, Varoquaux G, Gramfort A, et al. Scikit-learn: Machine learning in Python. the Journal of machine Learning research. 2011;12:2825–2830.

22. Hanley JA, McNeil BJ. The meaning and use of the area under a receiver operating characteristic (ROC) curve. Radiology. 1982;143(1):29–36.

23. Boyd K, Eng KH, Page CD. Area under the Precision-Recall Curve: Point Estimates and Confidence Intervals. In: Machine Learning and Knowledge Discovery in Databases. Springer Berlin Heidelberg; 2013:451–466.

24. Rodríguez-Pérez R, Bajorath J. Interpretation of Compound Activity Predictions from Complex Machine Learning Models Using Local Approximations and Shapley Values. J Med Chem. 2020;63(16):8761–8777.

25. Collins GS, Reitsma JB, Altman DG, Moons KGM. Transparent Reporting of a multivariable prediction model for Individual Prognosis Or Diagnosis (TRIPOD): The TRIPOD Statement. Ann Intern Med. 2015;162(1):55–63.

26. Akagi T, Nagata N, Miyazaki H, et al. Procalcitonin is not an independent predictor of 30-day mortality, albeit predicts pneumonia severity in patients with pneumonia acquired outside the hospital. BMC Geriatr. 2019;19(1):3.

27. Friedman JH. Stochastic gradient boosting. Comput Stat Data Anal. 2002;38(4):367–378.

28. Pourhoseingholi MA, Baghestani AR, Vahedi M. How to control confounding effects by statistical analysis. Gastroenterol Hepatol Bed Bench. 2012;5(2):79–83.

29. Bhasin A, Nam H, Yeh C, Lee J, Liebovitz D, Achenbach C. Is BMI Higher in Younger Patients with COVID-19? Association Between BMI and COVID-19 Hospitalization by Age. Obesity. 2020;28(10):1811–1814.

30. di Filippo L, Doga M, Frara S, Giustina A. Hypocalcemia in COVID-19: Prevalence, clinical significance and therapeutic implications. Rev Endocr Metab Disord. Published online April 13, 2021. doi:10.1007/s11154-021-09655-z

31. Zingmond DS, Parikh P, Louie R, et al. Improving Hospital Reporting of Patient Race and Ethnicity--Approaches to Data Auditing. Health Serv Res. 2015;50 Suppl 1:1372–1389.

